# Granular analysis reveals smart insufflation to be operationally more efficient and financially net positive compared to traditional insufflation for laparoscopic surgery

**DOI:** 10.1101/2023.04.11.23288414

**Authors:** Aazad Abbas, Imran Saleh, Graeme Hoit, Sam Park, Cari Whyne, Jay Toor

## Abstract

**Introduction:** Smart insufflation (SI) techniques relying on valve and membrane-free insufflation are increasing in usage. Although considerable literature exists demonstrating the benefits of SI on procedural ease and patient outcomes, there remains a paucity describing the financial impact of these devices. The purpose of this study was to determine the financial and efficiency impact of these devices on the operating room (OR) and inpatient wards of a hospital.

**Methods:** A discrete event simulation model representing a typical mid-sized North American hospital comparing SI to TI was generated. The National Surgical Quality Improvement (NSQIP) database from 2015 to 2019 was used to populate the model with data supplemented from literature. Outcomes included length of stay (LOS), duration of surgery (DOS), annual procedure volume, profit, return on investment (ROI), and gross profit margin (GPM).

**Results:** The operational parameters demonstrating favorability of SI to TI were DOS and LOS. DOS savings were 10-32 minutes/case while LOS savings were 0-3 days/case. Implementation of an SI led to an increase in annual throughput of 148 cases (12%). LOS decreased by 189 days (19%). This resulted in an increase in net profit of $104,675 per annum. The ROI of SI over TI device was >1000%.

**Conclusion:** Despite the initial financial investment being greater, the implementation of SI offsets these expenses and yields significant financial benefits. Our study demonstrates the financial benefits of SI over TI and illustrates how granular operational and financial analysis of technologies are essential to aid in sound healthcare procurement decision making.

## 1 Introduction

Despite being around for over a century, the use of laparoscopic surgery in modern clinical practice has rapidly increased over the last few decades^1^. This is due to the many advances in surgical technology, including live video feedback and abdominal insufflation techniques, as well as reduced postoperative pain, improved cosmetic results and reduced inpatient length of stay^1–5^. Insufflation involves the production of a pneumoperitoneum through the placement of CO2 gas into the peritoneal cavity, resulting in optimal visualization of the surgical field for surgical manipulation. As such, advances in insufflation technology, in particular “Smart” insufflation techniques, have tremendous potential to impact the field of laparoscopic surgery.

Traditional insufflation (TI) techniques involve the manual adjustment of the pressure of the pneumoperitoneum during the procedure, while smart insufflation (SI) techniques involve the use of low pressure and valve free access^6^. SI techniques are increasingly being utilized as they provide an immediate response time to even small changes in intra-abdominal pressure ^6–16^.These techniques allow for a more stable pneumoperitoneum during situations in which CO2 may escape or be eliminated from the cavity, such as during smoke evacuation or suction.

Although considerable literature exists to substantiate the benefits of SI regarding surgeon preference, procedural ease and improved patient outcomes, there remains a paucity in the literature on the granular financial impact of these devices ^7, 9, 10, 14–17^. This is an important question to address as these SI devices not only require a larger capital outlay than TI devices, but also increased disposable costs per case. However, they may potentially increase hospital efficiency to offset the initial and additional costs per case, leading to net financial positivity.

As such, the purpose of this study was to determine the financial and operational efficiency impact of the procurement of a novel SI device on a mid-sized hospital using comprehensive operational and financial modeling. We aimed to determine the financial and operational impact on the operating room (OR), duration of surgery (DOS) and inpatient length of stay (LOS) with respect to common procedures requiring insufflation.

## 2 Methods

### 2.1 Study Design

This was a retrospective database study utilizing a discrete-event simulation (DES) model representative of daily OR schedule and inpatient ward patient flow for gynecology procedures requiring insufflation at a mid-sized 500-bed academic-affiliated hospital performing approximately 10,000 procedures per year, 2321 number of which are gynecology, 1027 of which can be performed with a SI device. Research ethics approval from Mount Sinai Hospital were obtained before the commencement of this study.

Two scenarios were tested in the model: 1) using a TI device and 2) using a novel SI device. Ten thousand simulation replications were run and common random numbers were used when comparing scenarios to reduce the variability when comparing the outputs.^12^ A comparison was made between the two scenarios across key performance metrics namely duration of surgery, procedures completed per 8 hour window, annual capacity (number of procedures completed and patients managed per year), staffing requirements (with associated labour costs), length of stay, and inpatient versus outpatient ratio.

These results were then used as the inputs to a discounted cash flow (DCF) model. The DCF model was constructed in Excel using native formulae, with additional model input assumptions based on literature values as well as financial information provided by our accounting department. We assumed a 12-year time horizon based on estimated insufflator as provided by our hospital’s accounting department.

The cash flow in (CF) consisted of increased profit, difference in acquisition costs, and difference in disposable cost per surgery between TI and SI. The model then outputs key financial metrics including net present value (NPV), internal rate of return (IRR), return on investment (ROI), and gross profit margin (GPM).

### 2.2 Data Sources

Our hospital’s digital OR information system provided the list of gynecology procedures that could be performed with SI and their relative case-mixes. To allow for generalizable DOS and LOS results, the American College of Surgeons (ACS) National Surgical Quality Improvement (NSQIP) database was used to populate the DOS and LOS for each procedure. The accounting department provided all data regarding labour and capital costs. A literature review was performed to determine the impact of modern insufflators on procedure duration and hospital LOS for each procedure.

### 2.3 Discrete Event Simulation Model Construction

Patient flow through the OR and inpatient ward was first modeled as queueing networks, with ORs, post anesthesia care unit (PACU) beds and inpatient ward beds corresponding to servers, and patients to customers^18^. A discrete-event simulation model of the network was then developed in Python (Python Software Foundation, www.python.org) with its inputs using data from the OR workflow. A financial mapping, as described by Toor et al^19^, was performed to quantify the output of the models and compare the financial impact of both scenarios (TI vs SI). The model rests on two key assumptions. The first is that our hospital’s case-mix ratio is representative of similar sized institutions. The second is that inpatient ward time was assumed to be the LOS from the NSQIP database.

#### 2.3.1 Model Description

In the case of DOS simulations, scenarios with the TI and SI were identical except that every surgery in the SI is simulated to be a fixed amount of time shorter, representing time savings generated by SI use. Since the exact time saved by SI for the procedures selected is absent from the literature, several scenarios fixing the time saved at different values were tested. Cases were generated based on their probability of occurring at our institute, with DOS sampled with replacement from the NSQIP database. Cases were generated until their total duration exceeded a predetermined amount of time. Doing so allowed for a variable number of cases while minimizing the risk of cases running into overtime at any time step, avoiding the need to account for overtime costs. At our institution, the OR begins to incur overtime costs after 7.5 hours, and the average surgery is approximately 2 hours long, this predetermined amount of time was set to 5.5 hours in all scenarios. Each round of a scenario simulation was run for a total of 250-time steps, with each time step corresponding to a day in one OR. A time period of 250 days was assumed as there are 50 working weeks in the year (52 total weeks with 2 weeks of vacation) with 5 working days each week, resulting in 250 days. Since ORs do not share resources in the model, it was sufficient to model the patient flow through a single OR. Ten thousand rounds were simulated per scenario.

Similarly, the scenarios generated for inpatient days were also similar to each other, except that positive length of stays were decreased by a day in the scenario in which the SI was used. The types of surgeries and number of cases generated at each time step of the simulation were identical to as aforementioned. The time savings were incorporated into the generation of surgeries’ durations in the scenarios in which SI insufflators saved a positive amount of time in each surgery.

### 2.4 Discounted Cash Flow (DCF) Model Assumptions

The accounting department at our institution provided all financial inputs for the model. Cash outflows included an annual maintenance cost of $10,000 for TI and $15,000 for SI beginning year 1, disposable incremental costs of $75 for TI and $250 for SI, and an acquisition cost of $12,000 for TI and $22,000 for SI machines. Cost of capital used was healthcare industry standard 4.59%,^14^ with a time horizon of 12 years and a profit per case range of $500 to $1000. A base DCF model was constructed on the assumption of the midpoint of the range of the DOS and total hospital LOS savings per procedure, with a medium profit per case of $750. Sensitivity analysis was conducted by varying the DOS and LOS savings from 0 until the maximum reduction found in the literature, while varying the profit per case from $500 to $1000, to determine the difference in profit between the two scenarios. For each day reduction in total LOS produced by SI, it was assumed a similar laparoscopic procedure using TI was done.

### 2.5 Subgroup Analysis

Since the SI devices are specifically designed for complex procedures (e.g., hysterectomies), subgroup analysis was done with the above methods comparing all procedures with only complex gynecological procedures. These procedures were defined to be laparoscopic oophorectomy and total hysterectomies with and without salpingo-oophorectomy.

## 3 Results

Based on the literature review, the average time savings of smart insufflation was between 0- and 25-minutes surgical duration and between 0.5 and 2.5 days for inpatient length of stay reduction ^3, 9, 11–14, 20–23^. As such, duration of surgery reduction in the model ranged from 0 to 30 minutes and length of stay reduction from 0 to 3 days^3, 9, 11–14, 20–23^. Since the literature did not contain studies analyzing the time savings for gynecological procedures, we assumed the time savings would be similar for the currently available studies. The procedures included diagnostic laparoscopies, laparoscopic oophorectomy, laparoscopic resection of endometriosis, total laparoscopic hysterectomy with salpingo-oophorectomy and total laparoscopic hysterectomy without salpingo-oophorectomy. See Table 1 for relative institutional case mix, NSQIP DOS and LOS for included procedures.

**Table 1.** Case mix, NSQIP DOS, NSQIP LOS and CPT codes for the procedures used.

| Procedure name | Case Mix (%) * | Mean DOS (SD)† | Mean LOS (SD)† | CPT Codes |
| --- | --- | --- | --- | --- |
| Diagnostic Laparoscopy | 7.3 | 66.9 (42.4) | 4.9 (7.1) | 49320, 49321 |
| Laparoscopic Oophorectomy | 26.1 | 80.3 (51.6) | 2,0 (4.3) | 58661 |
| Laparoscopic resection of endometriosis | 14.3 | 89.4 (65.8) | 1.7 (2.7) | 58662 |
| Total laparoscopic hysterectomy with salpingo-oophorectomy | 33.1 | 126.8 (57.8) | 1.2 (1.9) | 58571, 58573 |
| Total laparoscopic hysterectomy without salpingo-oophorectomy | 19.1 | 132.4 (61.7) | 1.4 (2.2) | 58570, 58572 |

\*From our institutional dataset.
†From the NSQIP dataset.
DOS: duration of surgery, LOS: length of stay, NSQIP: National Surgical Quality Improvement Program, SD: standard deviation.

### 3.1 Scenario Comparison: TI vs SI

See Tables 2 and 3 for further details comparing the number of patients and throughput for all procedures and complex procedures. In terms of the number of procedures completed per year, the TI resulted in a total of 946 surgeries over a 250 day time frame. When varying the DOS time savings from 5 to 30 minutes when using the SI, the number of procedures completed per year ranged from 988 to 1292. This resulted in an increase in throughput ranging from 42 to 346 patients, a 4.4% to 36.6% increase in throughput. Similarly for complex procedures, 893 surgeries were done over 250 days, with SI increasing this to 931 to 1190 surgeries. Throughput increased from 38 to 297 patients, a 4.3% to 33.3% increase.

**Table 2.** Outputs of discrete event simulation (DES) models regarding number of procedures completed and total number of length of stay in terms of days per year for all procedures.

| Outcome Measure | TI | SI (5 min) | SI (10 min) | SI (15 min) | SI (20 min) | SI (25 min) | SI (30 min) |
| --- | --- | --- | --- | --- | --- | --- | --- |
| Number of Procedures Completed Per Year* | 946 | 988 | 1036 | 1089 | 1149 | 1215 | 1292 |
| Total number of inpatient days per annum for 1 day reduction per patient† | 957 | 598 | 627 | 658 | 695 | 738 | 782 |
| Total number of inpatient days per annum for 2 day reduction per patient† | 957 | 426 | 447 | 469 | 495 | 524 | 559 |
| Total number of inpatient days per annum for 3 day reduction per patient† | 957 | 343 | 359 | 378 | 397 | 421 | 447 |

\*At 250 days. †At 7.5 hours for every 250 days. TI: traditional insufflation, SI: smart insufflation.

**Table 3.** Outputs of discrete event simulation (DES) models regarding number of procedures completed and total number of length of stay in terms of days per year only for complex procedures (laparoscopic oophorectomy, total laparoscopic hysterectomy with and without endometriosis).

| Outcome Measure | TI | SI (5 min) | SI (10 min) | SI (15 min) | SI (20 min) | SI (25 min) | SI (30 min) |
| --- | --- | --- | --- | --- | --- | --- | --- |
| Number of Procedures Completed Per Year* | 893 | 931 | 971 | 1018 | 1069 | 1125 | 1190 |
| Total number of inpatient days per annum for 1 day reduction per patient† | 648 | 325 | 338 | 354 | 374 | 394 | 417 |
| Total number of inpatient days per annum for 2 day reduction per patient† | 648 | 195 | 204 | 213 | 224 | 237 | 250 |
| Total number of inpatient days per annum for 3 day reduction per patient† | 648 | 143 | 149 | 156 | 164 | 173 | 183 |

\*At 250 days.
†At 7.5 hours for every 250 days.
TI: traditional insufflation, SI: smart insufflation.

In terms of total number of days in the inpatient ward, the TI resulted in a total in patient LOS of 957 days. When varying the DOS time savings from 5 to 30 minutes when using the SI and assuming a 1 day LOS reduction for each patient when using an SI, the total number of inpatient LOS days ranged from 598 to 782 days, a reduction ranging from 175 to 359 days. Accordingly, the reduction in total inpatient length of stay ranged from 18.3% to 37.5%. Similarly, when assuming a 2 day LOS reduction for each patient when using an SI, the total number of inpatient LOS days ranged from 426 to 559, a reduction ranging from 398 to 531 days. Accordingly, the reduction in total inpatient length of stay ranged from 41.6% to 55.5%. Similarly, when assuming a 3 day LOS reduction for each patient when using an SI, the total number of inpatient LOS days ranged from 343 to 447, a reduction ranging from 510 to 614 days. Accordingly, the reduction in total inpatient length of stay ranged from 46.7% to 65.2%.

Similar results were noted for complex procedures, TI having 648 days on inpatient ward. The 1 day LOS reduction provided a decrease in inpatient days from 325 to 417 (231 and 323 days less), resulting in 35.6% to 49.5% reduction in total inpatient days. For a 2 day LOS reduction, total inpatient LOS ranged from 195 to 250 (398 to 453 days less), a 30.0% to 69.9% reduction. Again for 3 day LOS reduction, total inpatient LOS ranged from 143 to 183 (465 to 505 days less), a 71.8% to 77.9% reduction.

As expected, the total number of inpatient LOS increases when more time is saved for SI compared to TI; more procedures are performed, resulting in a larger throughput of patients in the system for all procedures in general and complex procedures only (Tables 2-3 and Figure 1).

**Figure 1.**
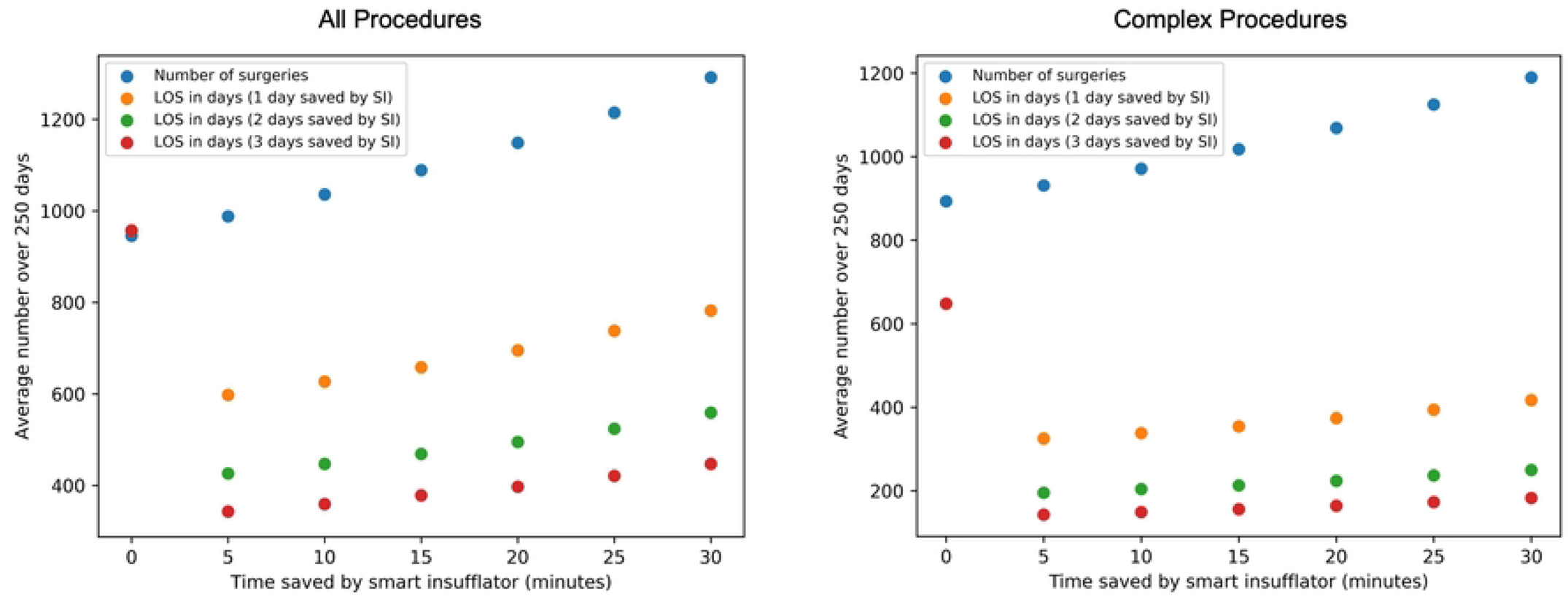
The average number of surgeries (blue) and length of stay (LOS) in days (orange) when the smart insufflator (SI) provides time savings ranging from 0 to 30 minutes for all procedures as well as complex procedures. 0 minutes corresponds to the traditional insufflator (TI).

### 3.2 Financial Analyses

Including all procedures, for the base DCF model, the profit from the TI was $638,550 and the profit from the SI was $746,325. Similarly, the GPM for the TI was 90.0% while the GPM for the SI was 71.7%. Regarding the difference between the two scenarios, the profit was $107,775, the NPV was $897,355.75, IRR was 294%, and the ROI was 1117%. For only the complex procedures, the profit for TI and SI was $602,775 and $707,450, respectively. GPM was similar at 90.0% for TI and 71.9% for SI. The profit difference was $104,675, NPV was $869,233.12, IRR was 285%, and ROI was 1081%.

Sensitivity table analysis of the profit difference between the TI and SI by varying the duration of surgery time savings (0 to 30 minutes), profit per case ($500-$1000) and length of stay savings (0 to 3 days) is shown in Figure 2 for all procedures and Figure 3 for complex procedures. For all procedures, length of stay savings of 0 days produced a positive profit difference only when the profit per case was above $900 for 25 minutes of DOS savings and above $750 for 30 minutes of DOS savings. For complex procedures only, length of stay savings of 0 days produced a positive profit difference only when the profit per case was above $950 for 25 minutes of DOS savings and above $800 for 30 minutes of DOS savings. For all procedures and length of stay savings of 1 day, as profit per case above and including $550 and DOS savings above and including 5 minutes resulted in a positive profit difference. For complex procedures and length of stay savings of 1 day, a positive profit difference was noted for profit per case above $500 for 20 minutes of DOS savings and for $550 for DOS savings above 5 minutes. For all and complex procedures for both length of stay savings above 2 and 3 days, all profit per case ranges ($500-$1000) and all DOS time savings resulted in a positive profit difference.

**Figure 2.**
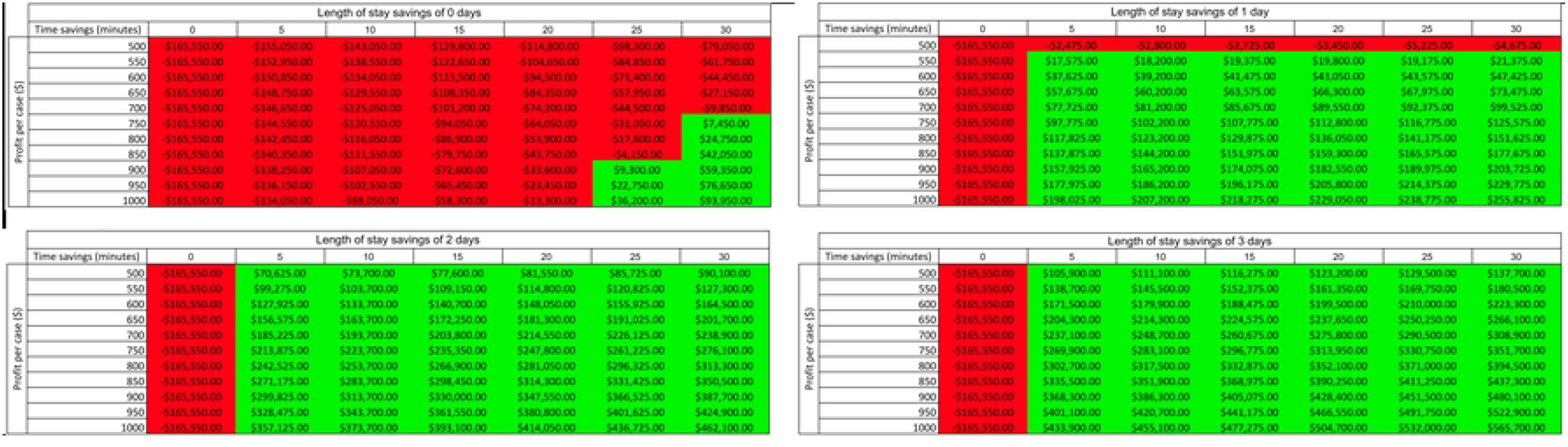
Sensitivity table for the difference in profit by varying time savings (0 to 30 minutes), profit per case ($500 to $1000), and length of stay savings (0 to 3 days) for all procedures.

**Figure 3.**
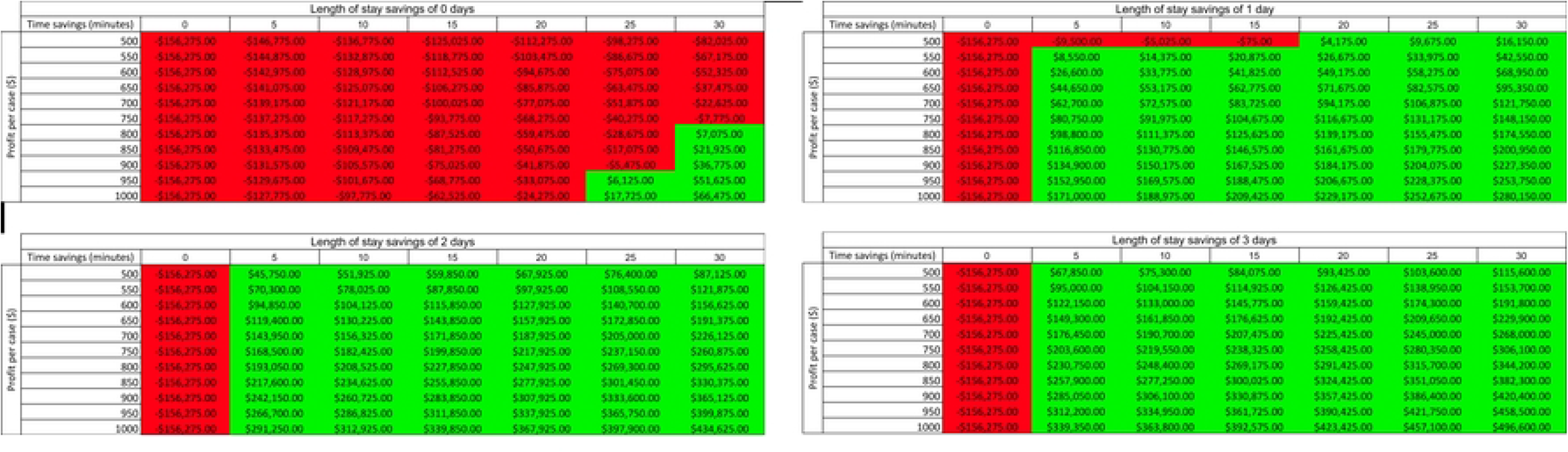
Sensitivity table for the difference in profit by varying time savings (0 to 30 minutes), profit per case ($500 to $1000), and length of stay savings (0 to 3 days) for complex procedures.

## 4 Discussion

Previous work examining the use of smart insufflation devices has focused on the clinical benefits and operational benefits when compared to traditional insufflation approaches ^11–16, 20–23^. However, there exists a gap in the granular financial quantification of the impact of these devices on a hospital level. This study was the first to quantify the financial impact on a hospital of using a smart insufflation (SI) device compared to a traditional insufflation device for gynecological procedures.

Using the parameters of our model, it was clear that SI increases the number of procedures done per year and decreases the total inpatient LOS at the same time. Of note, the more time saved for each procedure, the higher the throughput of patients per year and the larger the length of stay savings was. However, a key observation is that as the throughput of procedures per year increases, the total inpatient length of stay also increases as more procedures are done per year. This is key for hospitals as the more procedures done per year will result in more inpatient beds required, something they would need to consider from an implementation and operational perspective.

In terms of financial analysis, the base case clearly showed a larger profit for the SI as compared to the TI. The GPM was lower for SI as opposed to TI (71.7% versus 90.0%), which initially suggests TI to be favourable. This is due to ongoing disposable cost. Despite this, due to significantly increased throughput rate, SI was over one-hundred thousand dollars more profitable per annum. Another factor showing SI as more financially favourable than TI amortized over time was the high and positive NPV, IRR greater than cost of capital, and high ROI. It is important that hospitals consider these key financial metrics from a decision making perspective, especially when deciding to implement the use of a SI device.

Sensitivity analysis revealed important considerations. Firstly, it was clear that if the length of stay savings was 0, the SI would need to save at least 25 minutes from the procedure duration for each procedure in order to produce a profit greater than the TI. In addition, the profit per case would need to be at least $900 for the 25 minute time reduction and $750 for the 30 minute time reduction. However, as soon as a consistent length of stay reduction of at least 1 day is possible, the SI becomes greatly more profitable to the hospital with even a 5 minute savings in duration of surgery for each procedure (Figure 2). This clearly demonstrates that the SI devices need to provide efficiency in terms of duration of surgery and length of stay in order for them to be financially favourable to a hospital. Secondly, if the total inpatient length of stay savings provided by the use of SI devices are not replaced by similar procedures e.g. in this case the replacement procedures were those that can be done with TI devices, then the financial benefit of implementing an SI device is largely decreased. This is evidenced by the case where length of stay savings is 0, hence limiting the profitability of SI devices (Figure 2). Lastly, it is clear that the majority of the financial benefit of SI comes from the length of stay reductions it provides. The profit difference dramatically increases with each day reduction in length of stay. Institutions should keep this in mind when purchasing SI devices if they would like to reap the financial benefits of these devices.

This study presents with some limitations. Firstly, the length of stay savings provided by the SI were directly translated into additional procedures at the same profit margin. It may not be practically feasible for a hospital to replace each length of stay savings provided with such a procedure. They may be replaced with more expensive procedures, or not replaced at all, limiting the profitability of our scenario. Secondly, in reality the profit per case assumed in the financial analysis is dependent on each surgical procedure done and is not fixed at a particular value. As such, the real world returns on profit will largely vary according to the procedures done at each institution. However, it is challenging to account for this variability and our sensitivity analysis using a range of profit values is an attempt to account for this variability. Thirdly, the reduction in DOS and LOS provided by SI in the literature included gynecological and non-gynecological procedures, which was extrapolated and applied to the model. This speaks to the limited studies exclusively looking at the benefits of SI on gynecological procedures, something which future studies should expand on. Fourthly, the modeling in this study included simple procedures (e.g. diagnostic laparoscopy) and complex procedures (e.g. total hysterectomies) based on their relative case mixes to accurately represent a hospital’s gynecological service. However, SI devices are specifically designed to produce improved DOS and LOS savings compared to TI for complex procedures. As such, the savings demonstrated in this study could be substantially higher should only complex procedures use SI while simple procedures use TI. Lastly, we assumed our hospital’s case-mix ratio is largely representative of similar sized institutions. In reality, this is not the case and hospitals range in their relative case-mixes, even on a yearly basis. As such, this may limit the applicability of the exact profit values for each hospital. However, the approach demonstrated in this study may be applied to each institute to granularly quantify the financial effect on their hospital.

In conclusion, this study has demonstrated through DES operational modeling and DCF financial analysis that a novel smart insufflation device is financially more profitable than a traditional insufflation device on a mid-sized hospital. Classically, incremental costs per procedure have been a deterrent for procurement agents, as SI devices have higher incremental costs than TI devices. However, this study shows that procurement agents should attempt to quantify the entire operational and financial effect that using such devices would have on their hospital using metrics such as profit, NPV, IRR, and ROI. Future work would involve quantifying the real-world savings and financial benefits of implementing SI devices on an institutional level.

## Data Availability

Data used in this study can be found in this manuscript as well as the American Academy of Surgery (ACS) National Surgical Quality Improvement Program (NSQIP) database.

https://www.facs.org/quality-programs/data-and-registries/acs-nsqip/

## Notes

### Competing Interest Statement

The authors have declared no competing interest.

### Clinical Trial

NA

### Funding Statement

Research funding to conduct this study was provided by a research innovation grant provided by ConMed Corporation (#IRB 22-0113-C). These funds were used for student researcher remuneration, software, and manuscript dissemination. The funders had no role in study design, data collection and analysis, decision to publish, or preparation of the manuscript.

### Author Declarations

Mount Sinai Hospital Research Ethics Board Committee 700 University Avenue, 8th fl., Suite 8-600 Toronto, Ontario, Canada, M5G 1Z5 t: (416) 586-4875 f: (416) 586-4715 www.mtsinai.on.ca

